# The Protective Role of Belonging and Socioeconomic Status in Dropout Intent Among Minority Ethnic Students: A Mixed Methods Study

**DOI:** 10.64898/2026.06.12.26355506

**Authors:** Eleftheria Vaportzis, Maria Khan, Karisha Kimone George

**Affiliations:** Department of Psychology, University of Bradford, Bradford, UK; Department of Psychology, University of York, York, UK

**Author notes:** **Corresponding author details:** Eleftheria Vaportzis, Department of Psychology, University of Bradford, Bradford, BD7 1DP, UK.

**Keywords:** social integration, student retention, higher education equity, curriculum diversity

## Abstract

Improving minority ethnic student retention is a global higher education priority. This mixed-methods study investigated how institutional belonging and socioeconomic status interact to shape dropout intentions among minority university students in the UK (N = 182). Quantitative results revealed that perceived course difficulty and lower subjective socioeconomic status were the strongest predictors of dropout intent. While the interaction between socioeconomic status and difficulty was non-significant, qualitative accounts showed distinct structural vulnerabilities. Financial strain restricted social integration, turning socioeconomic disparities into campus isolation. Conversely, representative curricula, diverse peer networks, and stable cultural in-groups (e.g., religious affiliations, living in the parental home) functioned as essential psychological buffers against academic exhaustion and alienation. Universities must shift from transactional models to sustained structural equity to protect vulnerable student groups.

## Introduction

Improving student retention and fostering equitable educational experiences remain central priorities within higher education internationally, particularly as universities seek to widen participation among historically underrepresented groups (Crawford et al., 2024; Office for Students, 2024). Student withdrawal has substantial consequences for individuals, including reduced educational attainment, financial loss, and diminished career opportunities (Crawford et al., 2024). Although access has expanded, minority ethnic students continue to report inequitable academic and social experiences that can undermine their sense of belonging and increase dropout risks (Dost & Mazzoli Smith, 2023; Owusu-Agyeman, 2021). While previous research identifies belonging as a critical predictor of student engagement, wellbeing, and retention (Ahn & Davis, 2023; Dost & Mazzoli Smith, 2023), the mechanisms through which social, cultural, and economic factors interact to shape dropout intent among minority ethnic students remain insufficiently understood.

Sense of belonging refers to students’ perceptions of being accepted, valued, and included within their educational environment (Ahn & Davis, 2023; Gilani & Thomas, 2025). Bean and Eaton’s (2001) psychological model of student retention proposes that persistence is strongly driven by academic and social integration, alongside psychological adaptation. Thus, retention is driven by academic performance alongside subjective perceptions of institutional inclusion and connectedness (Arday, 2018).

Social Identity Theory (Tajfel et al., 2001) is a central theoretical framework to understanding sense of belongingness and student dropout intent. The theory suggests that individuals derive part of their self-concept from membership within valued social groups. Students are more likely to identify positively with their institution when they perceive that their cultural, ethnic, or social identities are recognised and represented within university spaces (Janke et al., 2024; Tajfel et al., 2001). Conversely, experiences of marginalisation, cultural disconnection, or racism weaken this identification and students may question whether they ‘fit’ within the higher education setting; this increases disengagement and withdrawal intentions (Janke et al., 2024).

At the institutional level, universities have increasingly emphasised inclusive curricula to foster belonging. Students who encounter diverse perspectives within reading lists, and classroom discussions report greater feelings of recognition, inclusion, and engagement (Barkas et al., 2022; Taff & Clifton, 2022). However, many minority ethnic students continue to report experiences of underrepresentation and cultural invisibility within course content (Denson & Bowman, 2017; Riedel et al., 2023).

Social integration serves as an important protective factor against student attrition (Abdul-Rahaman et al., 2023). Opportunities to develop authentic peer relationships and engage in extracurricular activities can strengthen students’ connection to the institution, foster emotional support networks and reduce feelings of isolation and withdrawal intentions (Abdul-Rahaman et al., 2023; De Sisto et al., 2022). Social Identity Theory suggests that membership of supportive peer groups can strengthen students’ sense of identity and belonging within the university community by fostering feelings of acceptance and group membership (Tajfel et al., 2001).

However, experiences of belonging are not uniform and are often shaped by intersecting demographic and socioeconomic factors. Financial hardship, for instance, has been linked to increased stress, reduced participation in university life, and lower levels of belonging (Bean & Eaton, 2001; Nguyen & Herron, 2021; Reid et al., 2020). Students who feel unable to participate in normative student activities may experience exclusion and reduced group membership. Consequently, lower socioeconomic status (SES) and financial disadvantage can drive dropout intent, both directly through practical barriers and indirectly by weakening belonging and institutional connectedness (Ahn & Davis, 2023). Beyond these social barriers, lower-SES students often face competing pressures from employment, commuting, caregiving, or limited resource access, further hindering academic engagement and persistence (Bean & Eaton, 2001). Similarly, for students with strong religious identities, campus environments that accommodate religious diversity foster greater belonging and psychological wellbeing (Owusu-Agyeman, 2021). Viewed through Social Identity Theory (Tajfel et al., 2001), religious affiliation offers group membership, shared values, and social connectedness. Because religious identity deeply intersects with ethnic and cultural identity for many minority ethnic students, institutional recognition of this diversity strengthens university identification and lowers withdrawal risk (Bean & Eaton, 2001; Owusu-Agyeman, 2021).

Despite these distinct threads, limited research explores how these intersecting demographic factors collectively shape dropout intent among minority ethnic students in UK higher education. Drawing upon theories of student retention and social identity, the present study therefore uses a mixed-methods approach to explore the factors associated with dropout intent among minority ethnic university students in the UK, focusing on the interplay between belonging, social integration, curricular representation, financial circumstances, and subjective SES. These factors were specifically chosen as they are among the most important dimensions of students’ psychological and social experiences in higher education and are particularly relevant for understanding dropout risk (Buizza et al., 2024). By combining quantitative relationships with qualitative accounts of students lived experiences, this research aims to provide evidence-informed insights for institutions seeking to create more inclusive and supportive learning environments to increase student success outcomes.

The study addressed three research questions:

- Which aspects of belonging and student experience are associated with dropout intent among minoritised ethnic students?
- Does subjective socioeconomic status predict dropout intent and perceptions of course difficulty?
- Does SES moderate the relationship between perceived course difficulty and dropout intent? Qualitatively, the study explored how students describe the factors shaping belonging and withdrawal intentions.

## Method

### Participants

Participants were required to be at least 18 years of age, currently enrolled as a university student, and identify as belonging to a minority ethnic group. Of the 197 total responses received, 182 were included in the analysis. Reasons for exclusion included not being a minority ethnic student (n = 11), not being a current student (n = 3), and having incomplete data (n = 1). Participants’ demographic information is presented in Table 1.

**Table 1.**
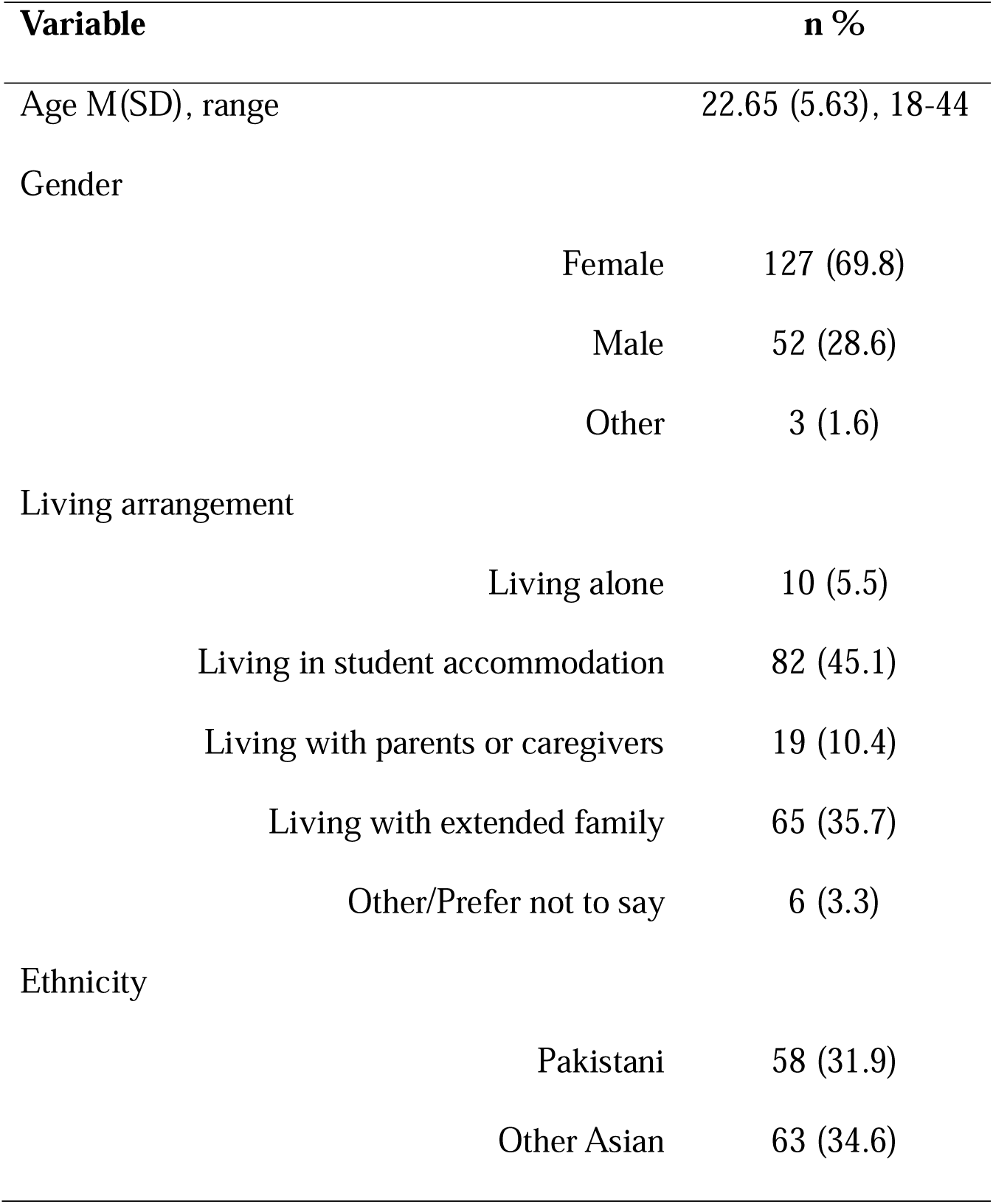
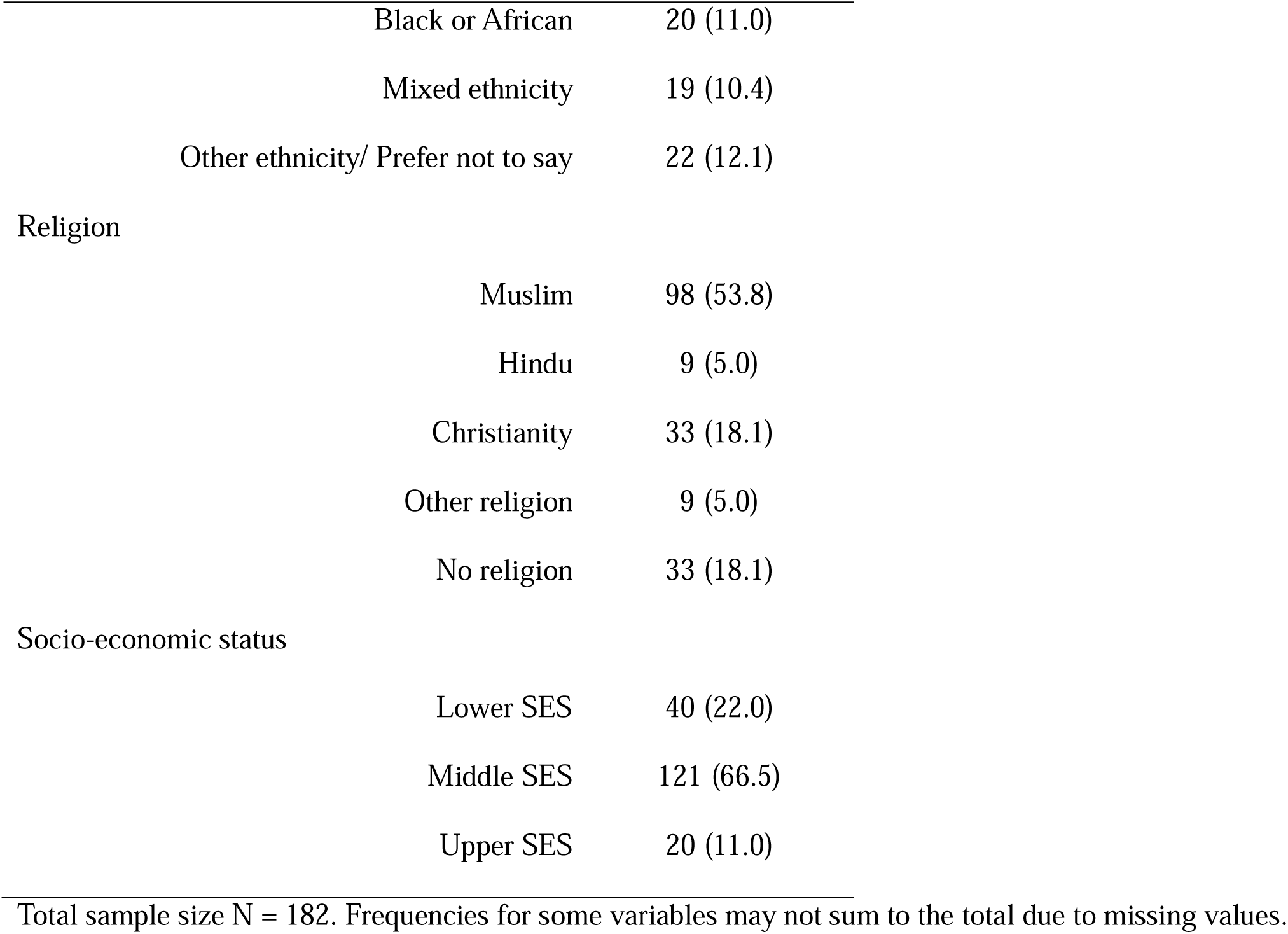
Sample demographic information.

### Recruitment

Participants were recruited via convenience sampling through student portals at the Universities of Bradford and York and UK student social media platforms. The survey was additionally promoted on social media platforms targeting UK university students. Data collection occurred across three phases between 2023 and 2025. The survey remained identical across all phases and data were aggregated into a single dataset for analysis (N = 182). No significant phase differences were identified in dropout intent or key variables; therefore data were pooled.

### Measures

#### Survey

A multi-dimensional survey assessed multiple aspects of student belonging and university experience. Survey items were developed from existing literature (see Table 2). Quantitative data were collected using statements measured on a 5-point Likert scale (1 = Strongly disagree, 5 = Strongly agree). Each scale item was accompanied by an open-ended question.

**Table 2.**
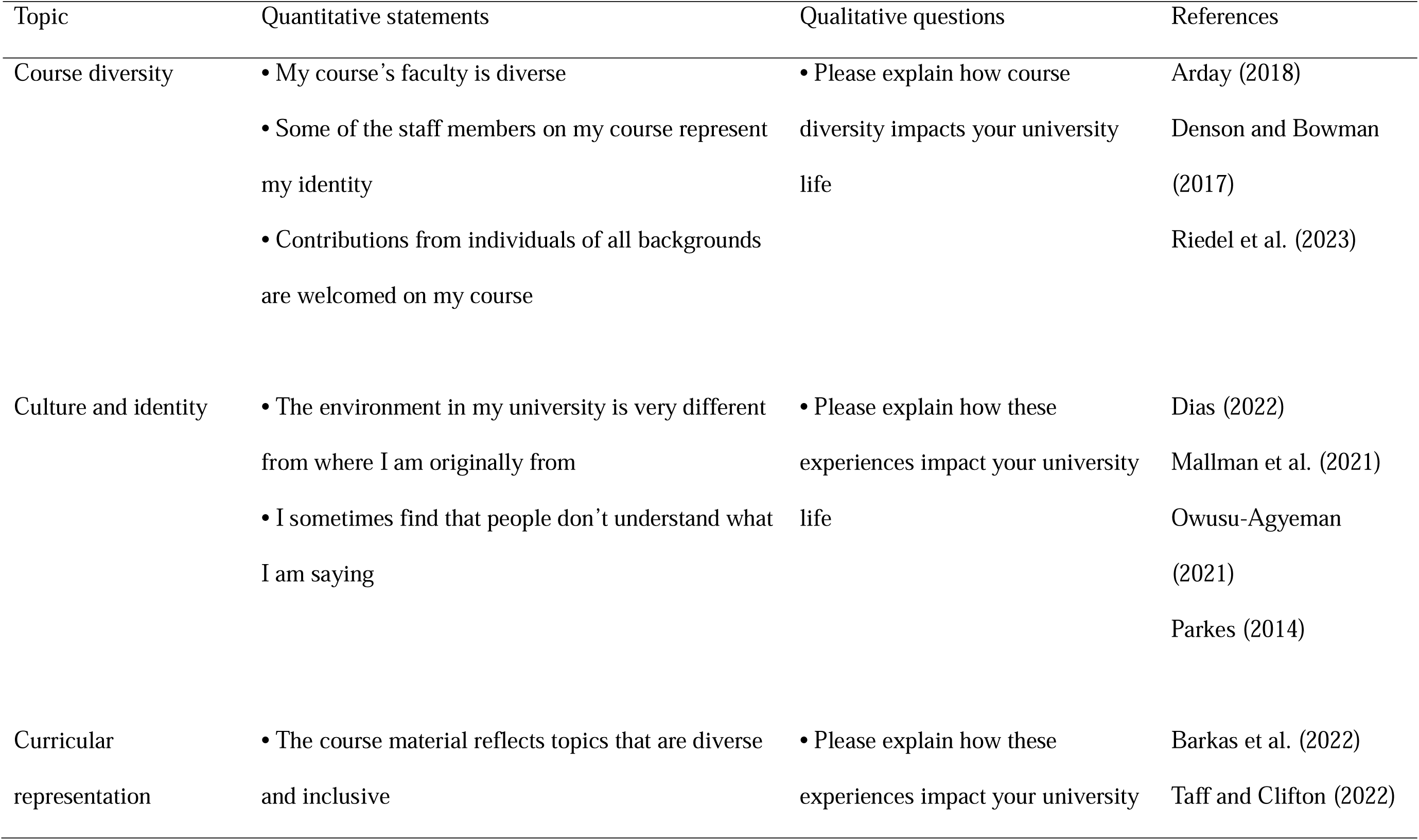
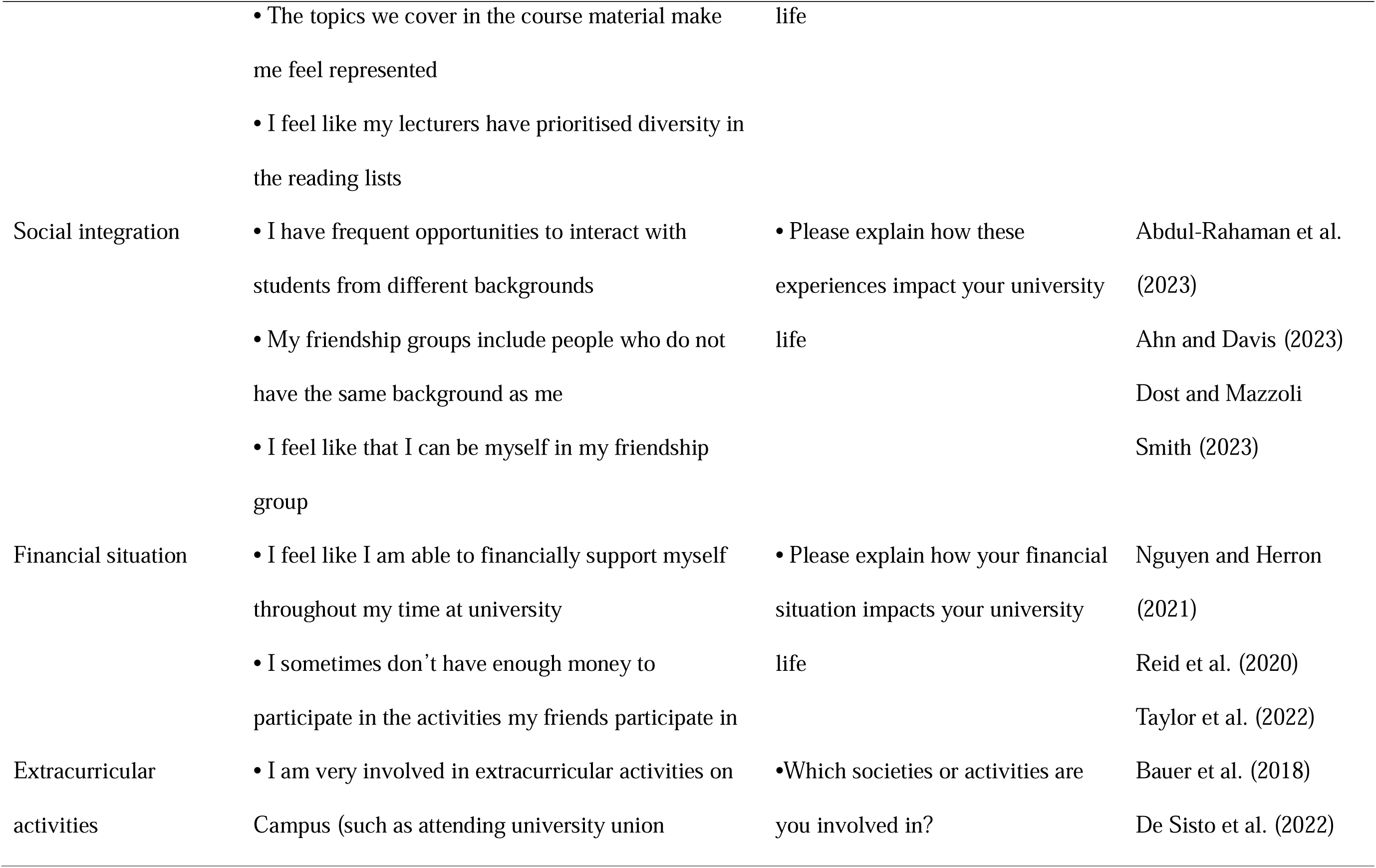
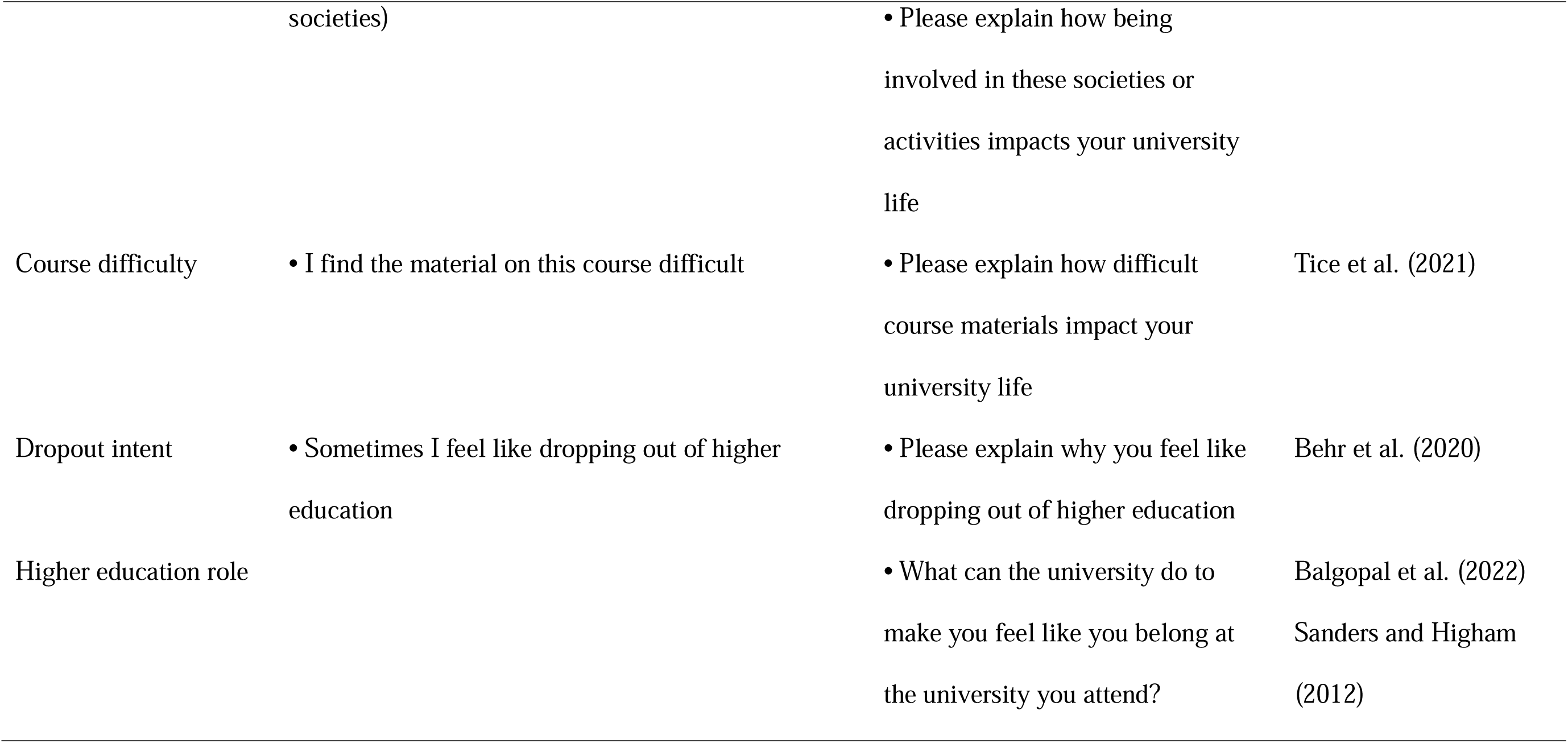
Summary of survey topics, items, and supporting literature.

Demographics included age, gender, ethnicity, religion, living arrangement and subjective socio-economic status (SES). SES was assessed using the MacArthur Scale of Subjective Social Status (Adler et al., 2000). Participants selected a rung on a 1–10 ladder representing UK social standing, which was aggregated into three tiers: Lower SES (Rung 1-3), Middle SES (Rung 4-6), and Upper SES (7-10).

### Procedure

Following electronic consent, participants completed the 15-minute online survey. All data were collected anonymously. Ethics approval has been granted by the Chair of the Humanities, Social and Health Sciences Research Ethics Panel at the University of Bradford on 7 July 2023 (EC27898). Ethics approval was also granted by the Chair of the Psychology Research Ethics panel at the University of York on 3 May 2024.

### Data analysis

A convergent mixed-methods design was adopted. Quantitative and qualitative data were collected simultaneously through the same survey instrument and integrated during interpretation.

### Quantitative analysis

Prior to analysis, Shapiro-Wilk tests indicated non-normal distributions across all scales (*p* < .001), while Levene’s test confirmed homogeneity of variance (*p* > .05). Spearman’s rank-order correlations (r_s_) were used due to non-normal distributions.

Group differences across SES, ethnicity, religion, and living situation were examined using one-way Analysis of Variance (ANOVA). To account for the non-normal distributions of the scales, all ANOVA and moderation analyses were evaluated using bootstrapping (5,000 resamples). ANOVA assumptions were considered acceptable and bootstrapping safeguarded against non-normality. Post hoc comparisons were performed with Tukey’s HSD tests. A two-way ANOVA and a moderation analysis (PROCESS macro, Model 1; Hayes, 2017) examined whether SES moderated the relationship between course difficulty and dropout intent.

A priori power analysis using G*Power (Faul et al., 2009) with α = .05 suggested that the sample had enough power (0.80) to detect a medium effect (f = 0.25) as 159 participants were required.

### Qualitative analysis

Data were analysed using reflexive thematic analysis (Braun & Clarke, 2006). Qualitative analyses were based on open-ended responses provided by survey participants across all question prompts. Responses ranged from brief comments to detailed narratives and generated a substantial qualitative dataset that enabled exploration of recurring patterns across students’ experiences. Following initial coding by the lead researcher (EV), two researchers (KKG and MK) acted as reflective critical peers independently reviewing a purposively selected subset of 20 information-rich responses. Discrepancies were explored through reflexive dialogue until a thematic framework was developed. The breadth and depth of participant responses were considered sufficient to achieve sufficient information power for the study aims. Code frequencies were calculated solely as descriptive signposts for mixed-methods integration. The researchers reflected on how their professional and cultural backgrounds may shape interpretation. Reflexive discussions were maintained throughout coding and theme development.

## Results

### Scale development and reliability

A Principal Component Analysis with Promax rotation supported the factorability of the 18-item survey (KMO = .78; Bartlett’s χ2 = 802.61, *p* < .001, see Supplementary Material). Prior to analysis, several items were reverse-scored so that higher scores consistently reflected greater levels of the named construct; most items demonstrated acceptable communalities (h^2^ > .40). Cronbach’s α confirmed four reliable scales: Curricular Representation: (α = .85; 3 items), Financial Support: (α = .70; 2 items), Course Diversity: (α = .65; 3 items), and Social Integration: (α = .60; 3 items). While internal consistencies for Course Diversity and Social Integration were modest, they demonstrated strong validity without redundancy, as the Mean Inter-Item Correlation fell within Clark and Watson’s (1995) ideal .15 to .50 range (i.e., Course Diversity = .39, Social Integration = .33).

### Correlational analysis

Spearman’s correlations identified the strongest positive relationship between dropout intent and course difficulty (r_s_ = .51, *p* < .001), followed by subjective SES (r_s_ = −.25, *p* < .001). Conversely, significant negative correlations were observed with social integration (r_s_ = −.30, *p* < .001), extracurricular involvement (r_s_ = −.23, *p* = .002), and age (r_s_ = −.19, *p* = .01). Additionally, a significant positive correlation was observed with financial situation (r_s_ = −.18, *p* = .01), while a strong positive relationship between course diversity and curricular representation (r_s_ = .60, *p* < .001; (see Table 3).

**Table 3.** Correlational analysis.

| Variables | *M* | *SD* | 1 | 2 | 3 | 4 | 5 | 6 | 7 | 8 | 9 | 10 |
| --- | --- | --- | --- | --- | --- | --- | --- | --- | --- | --- | --- | --- |
| 1. Course diversity | 8.15 | 2.39 | — | -.38*** | .60*** | .33*** | -.09 | .15* | -.19** | -.21** | .12 | -.02 |
| 2. Culture and identity | 5.40 | 1.81 |  | — | .23** | -.07 | .19*** | .10 | .16* | .14 | -.14 | .07 |
| 3. Curricular representation | 8.76 | 2.74 |  |  | — | -.17* | .26*** | -.11 | .15* | -.18* | -.24*** | .12 |
| 4. Social integration | 6.67 | 2.23 |  |  |  | — | -.18 | .05 | -.11 | -.30*** | .07 | .09 |
| 5. Financial situation | 5.82 | 1.17 |  |  |  |  | — | -.01 | .11 | -.18* | -.06 | .14 |
| 6. Extracurricular activities | 3.43 | 1.29 |  |  |  |  |  | — | -.12 | -.23** | .01 | -.20** |
| 7. Course difficulty | 1.76 | 0.76 |  |  |  |  |  |  | — | .51*** | -.05 | .16* |
| 8. Dropout intent | 3.11 | 1.31 |  |  |  |  |  |  |  | — | -.19* | -.25*** |
| 9. Age |  |  |  |  |  |  |  |  |  |  | — | -.02 |
| 10. SES |  |  |  |  |  |  |  |  |  |  |  | — |

Higher scores indicate greater levels of the construct measured. Scale scores were computed by summing constituent items. \* $p < .05$ , \*\* $p < .01$ .
SES = Socio-economic status.

### Differences in student experience by socioeconomic status and ethnicity

One-way ANOVAs revealed that subjective SES had a significant main effect on dropout intent, *F*(2, 179) = 4.48, *p* = .01, η2 = .05, and perceived course difficulty, *F*(2, 179) = 3.17, *p* = .04, η2 = .04. Post-hoc tests showed lower-SES students reported higher dropout intent than middle (*p* = .03) and upper-SES peers (*p* = .03), and found courses more difficult than upper-SES students (*p* = .04).

Ethnicity significantly affected culture and identity, *F*(4, 177) = 7.08, *p* < .001, η2 = .19, and finances, *F*(4, 177) = 5.05, *p* < .001, η2 = .20. Post-hoc tests showed Pakistani students scored higher on culture and identity than Black/African (*p* < .001) and mixed-ethnicity students (*p* = .003). Mixed-ethnicity students reported lower financial security than Pakistani (*p* = .01) and Black/African peers (*p* = .02).

### Secondary demographic comparisons

Secondary comparisons revealed main effects for culture and identity, *F*(4, 177) = 4.66, *p* = .001, η2 = .10, and financial situation, *F*(4, 177) = 3.90, *p* = .01, η2 = .08, where the Muslim students reported higher culture and identity than those with no religion (*p* = .01), and students living with parents reported higher scores than those in university accommodation (*p* < .001). Independent samples *t*-tests revealed that more mature students reported significantly lower levels of curricular representation, *t*(179) = 3.34, *p* = .001, and higher financial strain, *t*(76.77) = 2.33, *p* = .02, than younger students.

### Interaction effects of SES and course difficulty

A two-way ANOVA was conducted to examine the effects of subjective SES and course difficulty on dropout intent. Results revealed significant main effects for both course difficulty (F(2, 171) = 30.64, *p* < .001, η2 = .26) and SES tier (F(2, 171) = 4.25, *p* = .016, η2 = .05). The interaction between SES and difficulty was not significant (F(4, 171) = .55, *p* = .70), indicating that the relationship between course difficulty and dropout intent is consistent across different SES backgrounds (see Figure 1). However, conditional effects from a follow-up moderation model (R^2^ = .28, *p* < .001) indicated that the positive relationship between course difficulty and dropout intent was strongest for students with lower SES (effect = .92, *p* < .001), gradually weakening as subjective SES increased (effect = .65, *p* < .001).

**Figure 1.**
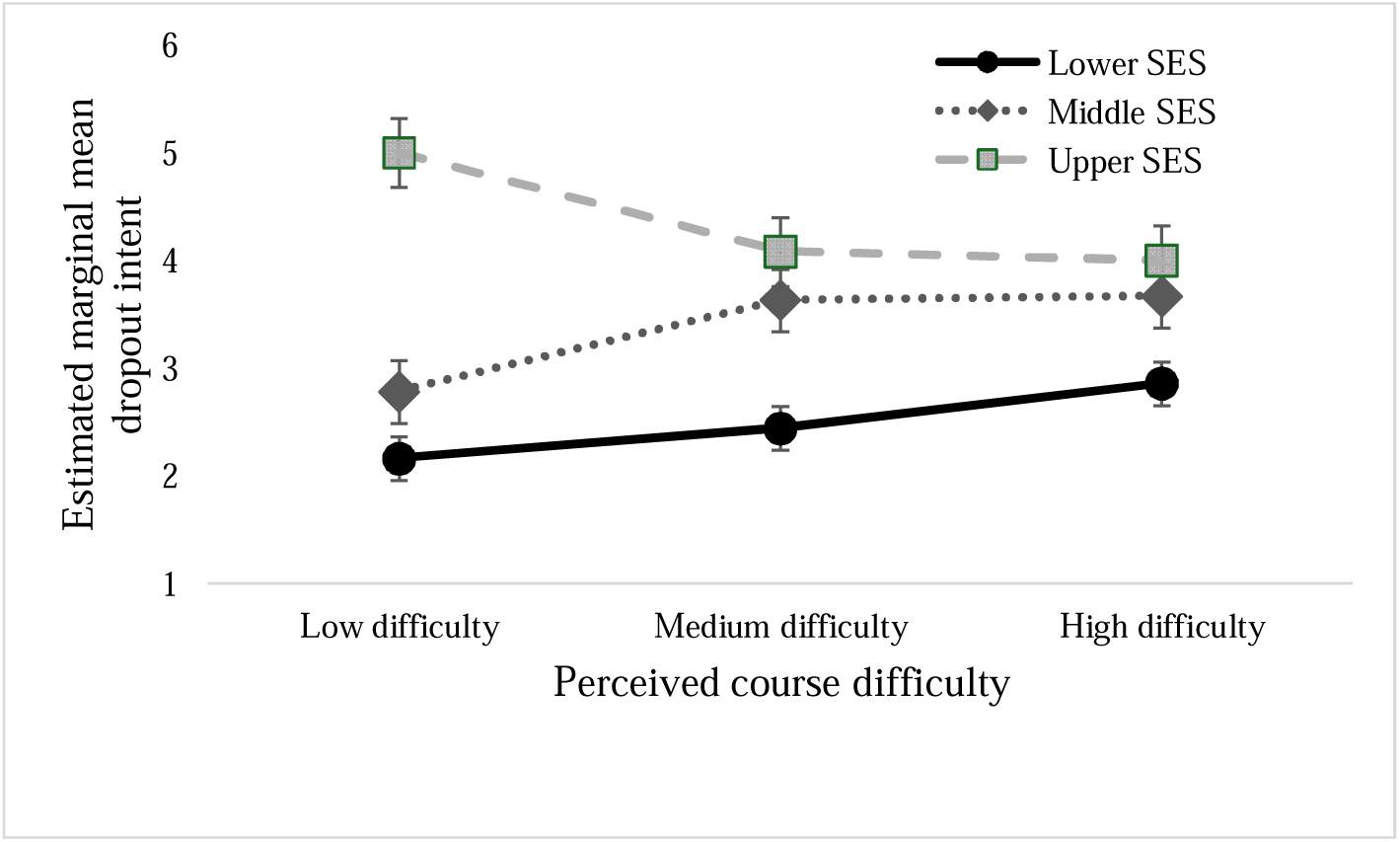
Estimated marginal means of dropout intent across socioeconomic status (SES) tiers and levels of perceived course difficulty. Error bars represent 95% confidence intervals. The results highlight significant main effects for both course difficulty (*p* < .001) and SES (*p* = .02), while the general trend of the plot lines illustrates a non-significant interaction (*p* = .70).

### Qualitative analysis

Reflexive thematic analysis of the open-ended responses captured the lived realities of minority ethnic students. Following a critical peer review process, the resulting dominant themes are summarised below by question topic including illustrative quotes. Qualitative responses were analysed by survey topic rather than PCA-derived factors to preserve contextual specificity.

### Course diversity

Student reflections indicated that representation within the academic environment strongly impacts psychological adaptation. Participants expressed that seeing staff and peers from matching backgrounds provides vital personal validation, helping them feel accepted and empowered to be themselves (“I feel like I belong in my course”), suggesting that demographic visibility actively validates their academic presence. Furthermore, structural representation within modules was associated with engagement and career aspirations (“It inspires [me] to achieve more”). A counter-narrative emerged around ‘Exclusion and Isolation’, which captured the psychological friction minority students navigate when representation is absent (“I felt like an outsider, as I had not seen any individuals with a similar background to mine in the faculty”), positioning an unrepresentative faculty functions as an alienating mechanism, reminding students of their marginalised status. Finally, some participants maintained a stance of ‘Diversity Neutrality’. These participants viewed demographic diversity as having little bearing on their day-to-day university trajectory (“Don’t really impact my university life”).

### Course materials

Participants reported contrasting experiential patterns regarding course difficulty. The first major theme, ‘Stress and Cognitive Load’, captured the negative emotional toll of workloads, with many describing difficult material as stressful and emotionally demanding (“I think I react to stress quite negatively, and when I do get stressed I’m not able to learn effectively”). The second major theme, ‘Academic Adaptation’, encapsulated the varied ways students successfully navigated these pressures. Within this adaptive framework, some participants viewed rigour positively, directly associating intellectual challenge with personal growth (“Difficult is a good thing, if it was easy I wouldn’t be learning anything”). Others described the material as highly manageable (“Studying has been easy so far”). This also manifested as self-discipline and persistence, with students emphasising an independent drive to increase their academic efforts when encountering difficult topics (“[…] when a material is harder, I just spend more studying instead of going out”). This reliance on isolation and increased labour suggests that academic persistence often demands an unfair trade-off with personal wellbeing.

### Curricular representation

Student narratives regarding reading lists and instructional materials revealed highly contrasting impacts on their university experience. The primary theme, ‘Inclusive Curricula’, captured a demand for diverse perspectives. Participants noted that representation within modules validates their presence in higher education and deepens their academic focus (“I feel more engaged and included in my studies when my lecturers prioritise diversity in the reading lists”). On the contrary, a prominent counter-narrative emerged around ‘Content Underrepresentation’, which captured the frustration and emotional detachment minority ethnic students felt when their lived realities were entirely absent from core texts (“I don’t feel represented in the literature that we study”). This absence fosters Eurocentric alienation, implicitly messaging to students that their communities have not contributed to their field of study. Finally, a distinct cluster of students maintained a stance of ‘Content Neutrality’ prioritising practical skill acquisition and technical learning over demographic diversification (“It’s a science based subject so it doesn’t really focus on people or types of people”).

### Culture and identity

Students described belonging as requiring both acceptance from others and validation of their own cultural identities. The primary theme, ‘Intercultural Fit’, captured protective spaces where diverse values allow minority ethnic students to establish psychological comfort and safety. Many participants felt fundamentally respected and accepted by peers and staff despite differences in background, beliefs, or identity (“I respect the fact that although I’m from a different background and have different beliefs, I’m respected regardless”). A counter-narrative emerged around ‘Cultural Alienation’, exposing the psychological strain students experience in less inclusive environments (“It’s hard when you have to be louder than others to be heard”). Several participants described experiences shaped by intersecting identities, particularly ethnicity, gender, and culture (“Being a Pakistani woman […] you are expected to act in a certain way and uphold certain values”). These intersections of identity create rigid behavioural scripts that students must navigate, penalizing authenticity and restricting social mobility.

### Social integration

Peer relationships acted as a key mechanism through which students developed a sense of belonging. The primary theme, ‘Intercultural Friendship Formation’, captured a strong student appreciation for cross-cultural contact, detailing how minority ethnic students actively build diverse social networks. Within this dimension, participants noted that developing close bonds outside their own ethnic or cultural groups facilitates a form of cultural immersion that expands their personal worldviews (“Having a diverse friendship group allows for cultural immersion from backgrounds I didn’t have knowledge on before”). On the contrary, ‘Inclusion Barriers’ outlined the communication gaps, cultural divisions, or religious differences that restrict comfortable peer interaction (“I have to be politically correct a lot”). This constant self-censorship reveals an underlying anxiety, where fear of social misalignment prevents genuine connection. Finally, a vital pattern was identified regarding ‘Social Safety and Belonging’, which emphasised the necessity of psychological comfort within a student’s wider university network, describing environments where individuals felt supported and free to be themselves (“Uni is made by my friends. I feel most at home there”).

### Financial situation

The financial realities of navigating higher education imposed distinct boundaries on social participation and academic focus. The primary theme, ‘Financial Comfort and Support’, highlighted how stable funding allows students to focus on their degrees rather than economic survival (“Being able to financially support myself reduces financial stress and allows me to focus more on my studies”). While some students reported complete self-sufficiency, many relied heavily on external systems, including student loans or parental assistance (“I’m lucky to be supported by my parents”). A major counter-narrative emerged around ‘Financial Constraints’, exposing the social isolation caused by budgetary constraints. Financial deficits frequently forced students to opt out of peer bonding experiences (“I often have to say no to social outings with friends because I simply cannot afford it”). To survive within the university environment, students deployed various financial coping strategies, such as part-time employment and lifestyle budgeting, which ultimately limited their broader student experience (“Sometimes I have to budget my money which limits the activities I can do”). Collectively, these narratives suggest that financial hardship affects not only material participation within university life, but also students’ perceived legitimacy and comfort within higher education environments.

### Extracurricular involvement

Extracurricular involvement functioned as a pathway to belonging for some students but remained inaccessible to others due to competing work and commuting demands. The primary theme, ‘Social Connection and Personal Development’, captured a broad spectrum of participation, highlighting how minority ethnic students navigate institutional activities. Participants expressed that involvement in campus clubs and societies serves as a vital pathway for expanding their social networks and fostering community connections (“It has helped me to come out of my shell, and get to know a lot of people from the university”). ‘Disengagement and Practical Barriers’, accounted for students who remained disconnected from extracurricular life and captured the practical barriers that actively prevent participation in university life. For some students, external responsibilities such as part-time employment or long commuting distances limited their ability to join social activities (“I commute and work part-time, so I don’t really have time to stay on campus for societies”).

### Dropout intent

The findings highlighted a critical equilibrium between navigating exhaustion and maintaining educational commitment. A major theme emerged around ‘Commitment and Persistence’, which illuminated the protective strategies students deploy to anchor themselves within higher education. Within this theme, participants framed their university attendance as a deliberate, highly valued choice focused on future mobility (“I think a degree will only open doors”). This dedication coexisted alongside ‘Burnout and Emotional Exhaustion’ which captured academic pressure as a major reason for considering withdrawal (“Sometimes there’s a massive build-up of pressure and it all seems quite worthless sometimes”). This sense of worthlessness reveals that academic exhaustion can trigger a deep existential toll, undermining a student’s sense of purpose. Finally, this psychological burden occasionally culminated in students ‘Questioning Degree Utility’, where systemic inequalities led students to view their higher education journey through a lens of futility (“Sometimes I don’t see a point in trying when I will never exceed those people who were born into privileged families no matter how hard I try”). This links dropout intent to perceived structural inequalities and limited opportunities. Importantly, dropout considerations were rarely framed as a lack of motivation. Instead, they reflected academic and financial pressure, cumulative emotional exhaustion, structural disadvantage, and uncertainty about future opportunities.

### Institutional recommendations for belonging

Students’ recommendations centred on three priorities: improving peer connection, increasing representation, and providing responsive academic support. While some participants expressed satisfaction, stating they felt integrated (“Nothing [to say here] as I feel my university gives me a sense of belonging”), others highlighted structured peer connection as a vital mechanism to foster community through formalised peer connection (“Implementing a peer buddy system during the first few weeks would make a massive difference”). This linked to requests for inclusive environments with greater staff diversity and representative curricula (“Maybe try to have teachers from different backgrounds and colours teach one subject”), placing the responsibility for change on the institution rather than the student. Finally, students emphasised academic support, calling for targeted educational scaffolding that accounts for diverse learner backgrounds (“Take into consideration that not everyone has the same educational background and limitations for international students”), demonstrating that equity requires addressing structural disparities.

## Discussion

Our findings provide a comprehensive, multi-dimensional view of the factors driving dropout intent among minority ethnic students. By synthesising quantitative analyses with qualitative narratives, this study highlights how institutional factors and a psychological sense of belonging converge to shape student retention outcomes. This study extends retention literature by demonstrating how intersecting structural inequalities, specifically SES vulnerability and institutional belonging, are psychologically experienced within everyday university life.

The primary synergy in our data revolves around course difficulty and SES. Quantitatively, course difficulty demonstrated the strongest positive association with dropout intent, while lower subjective SES significantly correlated with a higher desire to withdraw. However, because SES was measured subjectively, these findings suggest that perceived socioeconomic disadvantage may amplify the psychological burden associated with academic challenges. The qualitative data contextualise these trends by revealing the psychological mechanisms at play. When encountering academic difficulty, students experienced emotional tolls captured by the theme ‘Stress and Cognitive Load’. However, a clear divergence emerged: some students exhibited ‘Academic Adaptation’, reframing intellectual rigour as an opportunity for personal growth or increasing their self-discipline. These findings complement the quantitative results, which identified perceived course difficulty as a significant predictor of dropout intent, suggesting that academic challenge may be experienced either as a source of growth or as a contributor to emotional exhaustion depending on students’ available coping resources.

Although the moderation effect was non-significant, the pattern of findings suggested that academic challenge may be experienced more intensely amongst lower SES students. The qualitative findings help contextualise this pattern by showing that financial strain forces practical constraints. Students with lower financial security often juggle part-time employment and long commutes, leaving them trapped under ‘Disengagement and Practical Barriers’ that restrict their time to process difficult academic material or seek institutional help. Importantly, financial hardship was not experienced solely as an economic issue but as a barrier to social inclusion and participation in university life, directly aligning with these quantitative socioeconomic findings. Furthermore, mature minority ethnic students (25+ years) reported significantly higher financial strain and lower curricular representation than younger students. This reflects competing employment and familial responsibilities, alongside a curriculum that does not always acknowledge the experiences of older learners (Bean & Eaton, 2001). This highlights the need for inclusive curricula that intersect across both generation and culture, rather than treating minority ethnic students as an age-homogenous group.

Social integration and extracurricular involvement both demonstrated significant negative correlations with dropout intent, serving as key protective factors. These findings align closely with theories of student persistence which argue that academic and social integration are central to retention outcomes (Samoila & Vrabie, 2023). The present data further suggests that students who reported stronger peer relationships and greater involvement in university life expressed lower withdrawal intentions and greater resilience in the face of academic and financial stressors. Theoretically, this expands Bean and Eaton’s (2001) framework by demonstrating that academic and social integration are not parallel pathways, but interdependent tracks for minority ethnic students. Furthermore, it refines Social Identity Theory (Tajfel et al., 2001) by showing that institutional ‘fit’ is directly constrained by material, socioeconomic realities, rather than purely psychological perceptions. Narratives around ‘Intercultural Friendship Formation’ and ‘Social Safety and Belonging’ validate these statistics, demonstrating that diverse peer networks expand student worldviews and cultivate an essential sense of being ‘at home’. When academic pressures build, students frequently described these social networks as important sources of emotional support and connection. Conversely, communication barriers, cultural misunderstandings, and under-representative curricula left minority ethnic students feeling like ‘outsiders’, which appears to lead to emotional detachment and intentions to withdraw.

### Strengths, limitations, and future research

A primary methodological strength of this study lies in its integrated mixed-methods design, which allowed for cross-validation, using qualitative depth to directly unpack complex quantitative trends. To ensure interpretive depth and credibility, researchers engaged in collaborative critical peer review rather than using rigid codebooks. Additionally, the final sample size (N = 182) provided sufficient statistical power, exceeding the power analysis requirement needed to detect medium effect sizes within our ANOVA models.

Several limitations must be acknowledged alongside directions for future work. Participants were recruited via convenience sampling, which potentially limits generalisability across the wider UK sector. To address this, future research should replicate this mixed-methods framework across a more diverse range of cultural, geographical, and industrial educational sectors to test the boundary conditions of these findings. Furthermore, data collection occurred across three phases but remained cross-sectional in nature, meaning we cannot draw definitive causal lines regarding the longitudinal trajectory of dropout intent. Future scholarship should employ longitudinal tracking to monitor how the relationship between perceived course difficulty, financial stress, and dropout intent shifts across a student’s multi-year academic lifecycle. Another measurement limitation involves the internal consistency metrics of some of the subscales, which remained modest despite meeting exploratory criteria for newly developed instruments. Future refinement and validation of these scales across broader institutional contexts would strengthen confidence in their psychometric stability.

Demographic differences highlighted the importance of adopting an intersectional perspective when examining belonging and retention considering cultural, economic, religious, and social identities. Pakistani students reported significantly higher culture and identity scores than Black or mixed-ethnicity peers, while mixed-ethnicity students experienced lower financial security. Similarly, our findings showed that Muslim students and those living with parents reported significantly higher culture and identity scores highlighting the protective power of stable ‘in-groups’ (Tajfel et al., 2001). This directly echoes qualitative evidence from George et al. (under review), which demonstrates that more ethnically diverse institutional contexts remove the burden of protective identity management (’cloaking’), allowing minority ethnic students to express their cultural and religious identities authentically through supportive peer networks. For Muslim students, shared faith offers vital social connectedness that shields them from campus isolation. For commuting students, the parental home acts as a cultural anchor and safe domestic sphere away from alienating, Eurocentric university spaces. While commuting limits extracurricular engagement, living at home provides an emotional buffer that reinforces a secure sense of self. To expand on this, future research should explore intra-group differences rather than treating minority ethnic students as a homogenous population. Finally, while institutional interventions like peer mentoring are highly recommended based on student feedback, future studies should experimentally evaluate their objective efficacy on the actual retention rates of minority ethnic students to optimise their delivery.

### Practical recommendations

First, piloting structured peer mentoring during the initial weeks of term can accelerate intercultural friendship formation and build institutional trust. Second, departments should diversify curricula and reading lists, as culturally representative materials validate student presence and encourage academic engagement. Third, embedding asynchronous foundational resources, assessment rubrics, and scaffolded milestones within the curriculum practically supports lower-SES, international, and mature students facing financial or time constraints.

Finally, leadership must widen recruitment pipelines and implement legally compliant positive action frameworks (e.g., tie-break provisions) to diversify staff. This broadens equitable access for qualified underrepresented applicants, providing relatable identity models vital for reducing isolation. This can help broaden equitable access for qualified underrepresented applicants, providing the relatable identity models vital for reducing isolation.

## Conclusion

This study integrated quantitative metrics and qualitative lived experiences to untangle the drivers of dropout intent among minority ethnic students. The results demonstrate that while high course difficulty and lower socioeconomic standing act as primary vulnerabilities for student withdrawal, an institutional sense of belonging—driven by diverse curricula, financial security, and robust social integration—serves as a vital protective factor. To bridge the gap between systemic expectations and ground-level equity, universities must move past transactional education models and actively foster inclusive spaces where minority ethnic students feel structurally represented, socially safe, and academically supported. Ultimately, improving retention requires universities to replace symbolic inclusion with sustained structural practices that cultivate equity, representation, and safety.

## Data Availability

The data that support the findings of this study are available from the corresponding author, [EV], upon reasonable request.

## Acknowledgements

The authors would like to express their sincere gratitude to Aneekah Ashfaq for her invaluable assistance in compiling the participant materials, securing ethics approval, and coordinating the data collection process.

## Funding

This work was supported by the University of Bradford.

## Disclosure statement

The authors report there are no competing interests to declare.

## CRediT author statement

**Vaportzis:** Conceptualisation, Methodology, Formal analysis, Investigation, Writing - Original Draft, Writing - Review & Editing, Visualisation, Supervision, Project administration, Funding acquisition. **Khan:** Conceptualisation, Methodology, Formal analysis, Investigation, Writing - Original Draft, Writing - Review & Editing, Funding acquisition. **George:** Conceptualisation, Methodology, Formal analysis, Investigation, Writing - Review & Editing, Supervision, Project administration, Funding acquisition.

## Notes

### Competing Interest Statement

The authors have declared no competing interest.

### Author Declarations

Ethics approval has been granted by the Chair of the Humanities, Social and Health Sciences Research Ethics Panel at the University of Bradford on 7 July 2023 (EC27898). Ethics approval was also granted by the Chair of the Psychology Research Ethics panel at the University of York on 3 May 2024.

