## Supplementary Materials for "The Protective Role of Belonging and Socioeconomic Status in Dropout Intent Among Minority Ethnic Students: A Mixed Methods Study"

Table S1. Principal Component Analysis eigenvalues and variance explained

| Factor | Initial Eigenvalue  Total | % of Variance Explained | Cumulative % Explained |
| --- | --- | --- | --- |
| 1. Curricular Representation | 4.39 | 29.25 | 29.26 |
| 2. Course Diversity | 1.76 | 11.90 | 41.16 |
| 3.Social Integration | 1.46 | 9.76 | 50.92 |
| 4. Financial Support | 1.14 | 7.62 | 58.54 |

Extraction Method. Principal Components Analysis. KMO = .78. Bartlett's Test of Sphericity. χ^2^(105) = 802.61, p < .001.

Table S2. Rotated pattern matrix for the 15 student experience survey items

| Survey Scale Item | Component 1 | Component 2 | Component 3 | Component 4 |
| --- | --- | --- | --- | --- |
| The topics we cover in the course material make me feel represented | .91 | -.09 | -.06 | .01 |
| The course material reflects topics that are diverse and inclusive | .90 | -.23 | .03 | .07 |
| I feel like my lecturers have prioritised diversity in the reading lists | .85 | -.03 | -.03 | .04 |
| Some of the staff members on my course represent my identity | .55 | .25 | -.03 | -.06 |
| My course’s faculty is diverse | .42 | .29 | .20 | -.07 |
| Contributions from individuals of all backgrounds are welcomed | .42 | .27 | .37 | -.08 |
| I sometimes find that people don’t understand what I am saying | -.22 | .76 | .07 | .17 |
| The environment in my university is very different from where I am from | .09 | .68 | -.50 | -.11 |
| I feel like that I can be myself in my friendship group | -.09 | .53 | .43 | -.02 |
| Both staff and my peers accept that I may have different values to them | .21 | .39 | .04 | .01 |
| Some people on my course expect me to behave a certain way because of my identity | .35 | .37 | -.12 | .09 |
| I have frequent opportunities to interact with students from different backgrounds | -.04 | .07 | .83 | -.02 |
| My friendship groups include people who do not have the same background | .06 | -.12 | .75 | .02 |
| I feel like I am able to financially support myself throughout university | .08 | -.04 | .02 | .86 |
| I sometimes don’t have enough money to participate in activities | -.01 | .23 | -.03 | .80 |

Table S3. Item communalities and retention decisions from the principal component analysis

| Survey Scale Item | Extraction h^2^ | Decision |
| --- | --- | --- |
| The topics we cover in the course material make me feel represented | .77 | Retained (Curricular Representation) |
| The course material reflects topics that are diverse and inclusive | .73 | Retained (Curricular Representation) |
| I feel like my lecturers have prioritised diversity in the reading lists | .70 | Retained (Curricular Representation) |
| I have frequent opportunities to interact with students from diff. backgrounds | .69 | Retained (Social Integration) |
| I feel like I am able to financially support myself throughout university | .66 | Retained (Financial Support) |
| The environment in my university is very different from where I am from | .65 | Retained (Social Integration) |
| I sometimes don’t have enough money to participate in activities | .59 | Retained (Financial Support) |
| I sometimes find that people don’t understand what I am saying | .57 | Retained (Social Integration) |
| My friendship groups include people who do not have the same background | .56 | Retained (Social Integration) |
| Contributions from individuals of all backgrounds are welcomed | .55 | Retained (Course Diversity) |
| I feel like that I can be myself in my friendship group | .47 | Retained (Social Integration) |
| Some of the staff members on my course represent my identity | .46 | Retained (Course Diversity) |
| My course’s faculty is diverse | .43 | Retained (Course Diversity) |
| Some people on my course expect me to behave a certain way because of my identity | .39 | Omitted (h^2^ < .40) |
| Both staff and my peers accept that I may have different values to them | .27 | Omitted (h^2^ < .40) |
